# Empowering adults to manage their hearing loss: assessing the benefits of user-controlled, smartphone-connected hearing aids

**DOI:** 10.64898/2026.08.30.26361775

**Authors:** David W. Maidment, Alia Habib, Rachel Gomez, Claire Benton, Melanie A. Ferguson

**Author notes:** **Corresponding author:** David Maidment, PhD, E., T.

## Abstract

The availability of hearing aids that can connect wirelessly to smartphone technologies via Bluetooth has grown exponentially in recent years. However, there is limited evidence assessing the benefits of user-adjustability afforded by these devices. This study aimed to assess the benefits of smartphone-connected hearing aids and an accompanying application (or app) in new and existing hearing aid users. In this single-centre, prospective, observational study, 44 adult hearing aid users (14 new and 30 existing) were recruited. Participants were fitted bilaterally with smartphone-connected hearing aids that could be adjusted by the user via an app. Self-reported outcome measures were collected at fitting and after seven-weeks of using the device in everyday life. For both new and existing hearing aid users, significant improvements in social participation, hearing-related fatigue, and hearing aid benefit and satisfaction were found. For existing hearing aid users, all outcomes were significantly better for the smartphone-connected hearing aids plus app in comparison to their existing hearing aids that did not connect to a smartphone, all with moderate-to-large clinical effect sizes (*d*> .6). User-controllability via the app was identified as the key benefit, and most participants (68%) reported that the app met their needs ‘*extremely*’ or ‘*very well*’. These results suggest that, when used in conjunction with an app, smartphone-connected hearing aids can improve hearing outcomes due to greater user-controllability to improve listening. Thus, smartphone-connected hearing aids have the potential to facilitate patient-centred care, empowering the individual to successfully manage their hearing loss.

## INTRODUCTION

Smartphones that integrate the functionalities of a mobile (or cellular) phone and handheld computer have become a ubiquitous part of everyday life. It is estimated that 5.3 billion people worldwide (67% of the global population) own a mobile device, with approximately 75% of mobile connections operated on smartphones (GSMA Intelligence, 2022). Although often more synonymous with a younger demographic, the proportion of older adults that own a smartphone has grown exponentially. In the United Kingdom (UK), for example, smartphone ownership in young adults (16-24 years) remained unchanged at 96% in 2019 and 2021. In comparison, smartphone ownership in older adults (55+ years) increased from 52% in 2019 to 78% in 2021 (Ofcom, 2021). Given that smartphone adoption rates continue to rise in older adults, it is perhaps unsurprising that they now play a pivotal role in the management of age-associated, chronic health conditions (Cucciniello et al., 2021). Within the context of hearing loss, most leading manufactures have expanded their device portfolio to include hearing aids that connect to and can be adjusted via smartphone technologies, either by the user or remotely by a clinician. As a result of increasing market penetration, and in accordance with evidence-based practice, the benefits of user-adjustable smartphone-connected hearing aids compared to conventional hearing aids in adult aural rehabilitation should be evaluated.

Hearing aids are the most common management option for adults with mild to moderate hearing loss, and have been shown to improve listening abilities, as well as hearing- and general-health related quality of life (Ferguson et al., 2017). Despite this, adults with hearing loss often report that they continue to experience communication difficulties, particularly in the presence of background noise, which can lead to suboptimal- or non-use of hearing aids (Bennett et al., 2018). Typically, hearing aids are programmed and adjusted to an individual’s prescribed target gain by a hearing healthcare professional in the clinic using specialist equipment. The audiologist can also enable a volume control, as well as additional programmes, which can be manually operated by the user via a switch on the hearing aid or a wireless intermediary device, such as a remote control. This functionality is intended to allow the user to optimise their hearing aid settings in specific listening situations away from the clinic. However, previous studies have shown that hearing aid users often report experiencing problems making changes to their programmes and are unsure how and when to use them for optimal benefit (Bennett et al., 2020; Bennett et al., 2017; Goggins & Day, 2009). As a result, a high proportion of hearing aid users frequently return to clinic to have their hearing aid programmes re-explained or to request further adjustments (Goggins & Day, 2009). To address this, supplementary online resources, such as C2Hear (https://www.c2hearonline.com/) and m2Hear (https://www.nottingham.ac.uk/helm/dev-test/m2hear/), have been developed that can explain the use of programmes, as well as inform hearing aid users on further actions (e.g., communication tactics), minimising the need to return to the clinic (Ferguson et al., 2021; Ferguson, Maidment, Henshaw, & Gomez, 2019; Maidment et al., 2020).

The introduction of wireless streaming via Bluetooth, which allows the connection of compatible hearing aids to smartphone technologies, may provide an alternative to the provision of supplementary online resources. Specifically, smartphones provide a direct interface to hearing aids via an application (or app), which permits the user to adjust and customise their hearing aid programmes, as well as the volume, microphone directionality, and noise reduction algorithms, in any listening environment and without the need to visit the clinic. On this basis, smartphone-connected hearing aids have the potential to reduce the associated financial and opportunity costs for both the hearing aid user and audiology service provider. In addition, due to their advanced processing power, smartphones also enable additional functionalities, such as direct audio streaming (e.g. phone calls, music), geotagging for location-based configuration, use of the smartphone as a remote microphone system, connection to internet-based devices (or If This Then That, ITTT, networks), physical activity monitoring (e.g. step counter), fall detection, as well as remote programming by an audiologist (Kimball et al., 2018).

To explore the potential implementation of smartphone-connected hearing aids into clinical practice, several studies have assessed the attitudes of hearing healthcare professionals toward this technology (Kimball et al., 2018; Ng et al., 2017; Olson et al., 2022). In their qualitative collective case study undertaken in the United States of America (USA), Ng et al. (2017) found that clinicians reported improved rehabilitation practices as a consequence of fitting smartphone-connected hearing aids, as they dedicated more time getting to know the hearing difficulties that their patients were experiencing. Additionally, clinicians stated that linking smartphones with hearing aids reduced perceived stigma, as smartphone technologies were viewed as more socially acceptable (or ‘normalised’) in comparison to conventional hearing aids. Similarly, in a quantitative survey of USA-based audiologists, Kimball et al. (2018) found that clinicians were highly supportive of the integration of smartphone-connected hearing aids in adult aural rehabilitation. Nevertheless, while respondents advocated the use of smartphone apps by patients to make fine-tune adjustments and personalise their amplification settings, they were less supportive of allowing patients to make more permanent changes. A similar pattern of results has also been shown in a UK-based Delphi survey, a formalised methodology that seeks consensus amongst a panel of experts (Olson et al., 2022). Namely, Olson et al (2022) found that hearing healthcare professionals were generally accepting of smartphone-connected hearing devices because they perceived them to be easier and more convenient for patients to access.

Although there is a growing body of research showing that hearing healthcare providers are generally supportive of smartphone-connected hearing aids, there is a dearth of evidence assessing the benefits of these devices in adults living with hearing loss (Maidment et al., 2016, 2018). In a mixed-methods study, the everyday experiences of existing hearing aid users toward a range of smartphone-connected listening devices were evaluated (Maidment et al., 2019; Maidment & Ferguson, 2018). These devices included a smartphone-connected hearing aid, personal sound amplification product (PSAP), and a smartphone hearing aid app used with either wired earphones or a wireless hearable. Results from patient-reported outcomes showed that use, benefit, and satisfaction for smartphone-connected hearing aids were significantly better relative to conventional hearing aids, as well as all other devices studied (Maidment & Ferguson, 2018). Semi-structured interviews further revealed that the ability to make adjustments to hearing aid settings via a smartphone app resulted in a greater sense of autonomy, improved confidence, and increased social participation, which together empowered users to successfully manage their hearing loss (Maidment et al., 2019). This research has led to a series of studies investigating the underlying dimensions relating to empowerment in hearing healthcare. For example, Gotowiec et al. (2022) identified five dimensions of hearing-related empowerment including knowledge, skills and strategies, participation, self-efficacy, and control. This research has resulted in the development of two outcome measures, one for clinical use and another for research (Bennett et al., in review; Gotowiec et al., in review).

Nevertheless, it should be acknowledged that there are several caveats to the design of our previous mixed-methods study assessing smartphone-connected listening devices, including the recruitment of existing hearing aid users only. It could be argued that participants’ prior experiences of amplification may have introduced potential pre-exiting biases, limiting the generalisability of the study findings. In addition, the devices were trialled in the real-world for a relatively short time of two-weeks. This may have been insufficient for users to fully acclimatise to new device settings, which can take several weeks to achieve (Giroud et al., 2017). On this basis, we have undertaken a further mixed-methods study in both new and existing hearing aid users, whereby the devices were used for a longer time of seven-weeks. The qualitative component explored users’ perspectives toward the barriers and facilitators of using the smartphone-connected hearing aids (Gomez et al., 2022), which was underpinned by the capability, opportunity, motivation, and behaviour (COM-B) model and Theoretical Domains Framework (Michie et al., 2014). A key advantage identified of smartphone-connected hearing aids was that the accompany app improved knowledge of hearing aid controls and gave the user greater autonomy and control to fine-tune their device to meet their individual needs (capability). By being able to control the sound quality, participants were more likely to participate in conversations, and stigma was reduced because smartphones were viewed as ubiquitous (opportunity). For beliefs about capabilities (motivation), empowerment was as a key theme, as participants reported they could control and use their listening devices how and when they wanted, increasing self-management. To overcome barriers of perceived poor digital literacy skills and confidence in using smartphone technologies, Gomez et al (2022) also endorsed specific behaviour change techniques that could be incorporated into future interventions, including enablement, goal setting, reframing perceptions towards technology, and addressing educational needs.

In the current paper, the quantitative component of this more recent mixed-methods study is presented, which assessed the benefits, or otherwise, of user-adjustability afforded by smartphone-connected hearing aids and an accompanying app. Specifically, we report the extent to which smartphone-connected hearing aids plus app can impact self-report and behavioural measures of social participation, hearing-related fatigue, quality of listening, and hearing aid outcomes (i.e., use, benefit, residual disability, satisfaction).

## METHOD

### Study Population

Participants were recruited from the Adult Audiology Service at Nottingham University Hospitals National Health Service (NHS) Trust. Inclusion criteria were: (i) adults aged ≥18 years; (ii) symmetrical (no more than 20 dB HL differences across ears at octave frequencies 0.5-4kHz) sensorineural mild to severe hearing loss in N2, N3, N4, N5, S2 and S3 categories (Bisgaard et al., 2010); (iii) never used hearing aids (new user), or been using hearing aids for more than six-months (existing user); (iii) personally owned an iPhone 5 or higher with IOS ≥10 for compatibility with the smartphone app, and used for functions that exceed calling and writing SMS text messages; (iv) willing to use smartphone-connected hearing aids and accompanying app during the seven-week study period; and (v) have a good understanding of the English language in order to understand the intervention content. Exclusion criteria were: (i) disturbing tinnitus; (ii) contraindications against wearing hearing aids (e.g., ear disease, motor impairment); and (iii) unable to complete outcome measures unassisted due to cognitive decline or dementia assessed via self- or familial-report.

### Study Design

The design was a prospective, observational cohort study assessing the benefits of smartphone-connected hearing aids plus app when used by new and existing users in their everyday lives for seven-weeks. The study was prospectively registered on September 17^th^ 2018, at ClinicalTrials.gov (NCT03674086).

### Procedure

All patients who had been referred to the Adult Audiology Service in Nottingham, UK for an initial hearing assessment appointment or a hearing reassessment appointment were posted a study information pack containing an invitation letter, information sheet, reply sheet, and pre-paid envelope to return the necessary documents if they wished to take part in the study. In addition, at the end of their clinical appointment, patients were also asked by their audiologist if they would like to take part in the study. Those interested were referred to a member of the research team present in a separate clinic room. Details of the study were explained in the participant information sheet, as well as verbally by a member of the research team who also confirmed study eligibility and obtained informed written consent (see also, Figure 1).

**Figure 1.**
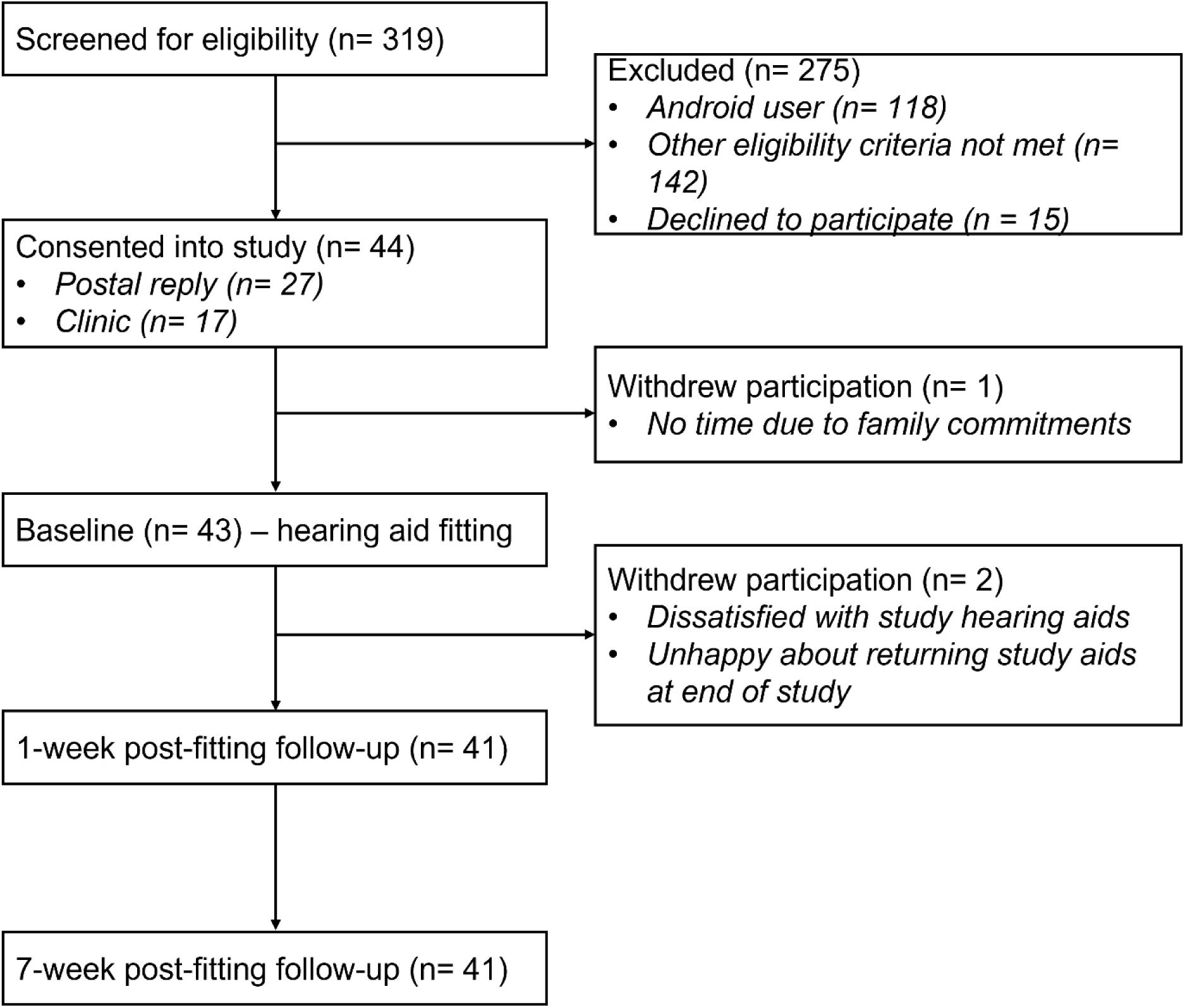
Flow diagram showing progression of participants through each phase of the study.

Post-consent, ear impressions were taken where necessary, and participants were invited to attend a baseline study session at the NIHR Nottingham Biomedical Research Centre (BRC). During this session, participants were fitted with bilateral smartphone-connected hearing aids by a qualified research audiologist (AH, RG). Once fitted, the audiologist ensured that the participants smartphone could successfully connect to the hearing aids via Bluetooth. The accompanying app was downloaded and full instructions on how to use all the app’s features was given. Participants also had the opportunity to practice adjusting the hearing aids using the app. A second visit was arranged one-week post-fitting, where any issues relating to the physical fit of the hearing aids were resolved, as well as changes to the gain if requested. A third follow-up visit was completed seven-weeks post-fitting, where the smartphone-connected hearing aids were returned and participants fitted with conventional NHS hearing aids in accordance with local protocols and national guidelines (British Society of Audiology, 2011).

All participants received a nominal inconvenience allowance and reasonable travel expenses for attending each study visit at the NIHR Nottingham BRC. The study was approved by the UK National Health Service (NHS) Health Research Authority, South Central – Berkshire B Research Ethics Committee and Nottingham University Hospitals NHS Trust Research and Innovation department.

### Study Interventions

*Smartphone-connected hearing aids*. Phonak Audeo B90-Direct hearing aids were fitted via NOAHlink Wireless to the Phonak Digital Adaptive prescriptive formula using pure-tone air- and bone-conduction thresholds from the participant’s latest clinical examination and were verified using in-situ audiometry via the Phonak Target 5.2 fitting software. Hearing aids were fitted with receiver-in-canal (RIC) custom-moulded shell (cShell) (n= 33), or slim-tube dome (n= 11) couplings based on clinical need and participant preference. All hearing aid fittings were undertaken in a sound-treated booth at the NIHR Nottingham BRC.

*Smartphone app*. Phonak Audeo B90-Direct hearing aids wirelessly connected to a smartphone app via Bluetooth. Specific features of the app included:

i. *Sound modifiers*. Enabled the user to control different sound settings, including overall gain (for each hearing aid separately or both), gain across three-channels (bass, middle, treble), as well as noise reduction, microphone directionality, and compression.
ii. *Factory pre-sets.* Allowed the user to select situation-specific programmes, such as restaurant, television, or music. Pre-sets were also available for quick changes to overall gain, such as *more bass* vs. *more treble*.
iii. *Custom programmes*. The user could create and save customised programmes using the different sound modifiers and factory pre-sets specified, which could then be saved within the app for future use. The app did not allow participants to adjust the maximum power output (MPO). Only limited ranges for adjustments were accessible based on the fitting ranges set by the audiologist.

### Study Measures

Outcome measures were administered face-to-face at baseline (i.e., smartphone-connected hearing aid fitting), as well as one- and seven-weeks post-fitting (Figure 1). On completion of the quantitative study, a sub-sample of participants (n= 8) also took part in one of two focus groups, the results of which are reported elsewhere (Gomez et al., 2022).

#### Clinical measures

The following measures were completed before smartphone-connected hearing aid fitting (i.e., baseline) only:

##### Hearing sensitivity

During participants’ hearing (re)assessment appointment, pure-tone air conduction thresholds were measured at octave frequencies (0.25 to 8kHz) for each ear, and bone-conduction thresholds as required (0.5 to 4kHz), following the procedures recommended by the British Society of Audiology (2011).

##### Montreal Cognitive Assessment (MoCA)

(Nasreddine et al., 2005). A validated screening assessment for mild cognitive impairment assessing several domains including short-term memory, visuospatial abilities, executive function, attention, language abilities, and orientation. Total scores range from zero to 30 points, with higher scores indicating better cognitive function. A score ≥26 is considered ‘normal’.

#### Validated self-report measures

The following outcome measures were completed by both new and existing hearing aid users at baseline and at the seven-week follow-up unless otherwise stated.

##### Glasgow Hearing Aid Benefit Profile (GHABP)

(Gatehouse, 1999). Completed by new hearing aid users only. Assesses hearing disability (activity limitations) and handicap (participation restrictions) (part I), as well as hearing aid use, benefit, residual disability, and satisfaction (part II) across four pre-defined listening situations. Each subscale is measured on a five-point scale. Part I was administered at baseline and part II at the seven-week post-fitting follow-up.

##### Glasgow Hearing Aid Difference Profile (GHADiffP)

(Gatehouse, 1999). Completed by existing hearing aid users only. Assesses use and residual disability with ‘old’ (i.e., conventional) hearing aids (part I) and use and residual disability with ‘new’ (i.e., smartphone-connected) hearing aids, as well as the difference in benefit and satisfaction between ‘old’ and ‘new’ hearing aids (part II). Part I was administered at baseline and part II at the seven-week follow-up.

##### Hearing Handicap Inventory for the Elderly (HHIE)

(Ventry & Weinstein, 1982). Assesses the emotional (13 items) and social/situational (12 items) impact of hearing loss using a three-point scale (4= *Yes*; 2= *Sometimes*; 0= *No*). Higher scores indicate greater impact of hearing loss.

##### Vanderbilt Fatigue Scale for Adults with Hearing Loss (VFS-AHL)

(Hornsby et al., 2021). Consists of the following four sub-scales, each with 10 items: cognitive, physical, emotional, and social listening-related fatigue. Each item is rated on a five-point scale. For items one to 31, participants are asked how often they experience or react in a certain way, in a given situation (0= *Never/almost never*; 4= *Almost always/always*). For items 32 to 40, participants are asked how much they agreed, or disagreed, with a particular statement (0= *Strongly disagree*; 4= *Strongly agree*).

##### Device Orientated Subjective Outcome Scale (DOSO)

(Cox et al., 2014). Assesses subjective opinions of hearing aids. Comprises of 40 items and produces scores for six subscales: speech cues (14 items), listening effort (10 items), pleasantness (four items), quietness (five items), convenience (four items), and use (three items). Each subscale is accompanied by a seven-point scale (A= *Not at all*; G= *Tremendously*), except for use, which is accompanied by a five-point scale. Completed at baseline for existing hearing aids users with respect to their conventional NHS hearing aids, as well as at the one- and seven-week follow-up sessions in respect to the smartphone-connected hearing aids.

##### Auditory Lifestyle and Demands Questionnaire (ALDQ)

(Gatehouse et al., 1999). Contained 24 listening situations (Supplemental Materials). For each situation, participants were asked how often they find themselves in that situation (1= *very rarely*, 2= *sometimes*, or 3= *often*), and how important it is for them to be able to hear in that situation in their everyday life (1= *very little*, 2= *some importance*, or 3= *very important*). Completed at the one-week follow-up session only.

#### Non-validated self-report measures

##### Initial Hearing Aid Preferences Questionnaire (IHAPQ)

Ten items (Supplemental Materials) assessing preferences concerning hearing aid design and use. Each was ranked on a response scale from one (*least important*) to 10 (*most important*). Completed at baseline only.

##### Clarity and Comfort Questionnaire (CCQ)

Comprised four items assessing individual preferences regarding sound quality from hearing aids on a two-point response scale (Supplemental Materials). Completed at the one-week follow-up session only.

##### Hearing Tasks Diary

Participants were required to compare the activation of specific gain settings (*more sound*, *more base*, *more treble*, *clarity*, *comfort*) for different factory pre-sets (*TV, restaurant, music, car*) within the app in specified listening situations (e.g., *watching a film, listening to music*), rating their preference on a five-point scale (−2= gain setting *‘on’ sounds much better*; -1= ‘*on’ sounds slightly better*; 0= ‘*on’ sounds the same as ‘off’*; 1= ‘*off’ sounds slightly better*; 2= ‘*off’ sounds much better*). Participants were instructed to complete the diary throughout the seven-week, home-based study period.

##### Feedback survey

Consisted of closed- and open-ended questions to assess participants’ views concerning the usability of the smartphone-connected hearing aids and app in the real-world (Supplemental Materials). Completed remotely online during the seven-week, home-based trial. In a slight deviation to the pre-registered protocol, due to low adherence the questionnaire was not completed at weeks two and six as originally intended. Instead, the feedback questionnaire was completed by participants on one occasion between weeks two and six.

#### Behavioural measures

##### Bamford-Kowal-Bench Speech-in-Noise (BKB-SIN) test

(Bench et al., 1979). Consists of 21 lists, each containing 16 phonetically balanced sentences (e.g., *the clown had a funny face*) with three or four target words scored per sentence (e.g., *clown*, *funny*, *face*). Completed in a sound-attenuated room, participants were seated in the centre of a five-loudspeaker array of radius 1.2 metres. Speech was presented at 0° azimuth and multi-talker babble noise was spatially separated at 45°, 135°, 225° and 315° azimuths. Using an adaptive procedure (MacLeod & Summerfield, 1990), sentence list one was first presented to estimate the 50% aided speech reception threshold (SRT) using the ‘automatic’ hearing aid factory pre-set. The speech level was then fixed at +3 dB SPL above the estimated SRT and performance tested for the following three hearing aid factory pre-set conditions: (i) restaurant/neutral; (ii) restaurant/clarity; and (iii) custom pre-set based on restaurant/neutral. Two sentence lists (excluding list one) were selected at random and administered for each test condition. The total number of correct keywords identified were recorded for each list presented. On completion of each test condition, participants were also asked to rank their perceived listening effort on a visual analogue scale, ranging from one (*No Effort*) to 13 (*Extreme Effort*). Completed by all participants at the one- and seven-week follow-up.

*Hearing aid use* (average hours/day) using datalogging integral to the smartphone-connected hearing aid was downloaded for the period between baseline and one-week post-fitting, as well as between one- and seven-weeks post-fitting.

### Statistical Analysis

#### Validated self-report measures

Prior to analysis, it was confirmed that measures did not violate the assumptions of normality or homogeneity of variance. For the GHABP (new users) and GHADiffP (existing users), differences between part I and part II subscales were examined using a paired-samples t-test. Similarly, for the HHIE and VFS-AHL, the difference between study sessions (i.e., baseline and seven-week follow-up) for new and existing hearing aid users was assessed using separate paired samples t-tests. For existing hearing aid users only, the difference between conventional hearing aids at baseline and the smartphone-connected hearing aids at both the one- and seven-week follow-up for the DOSO was examined using repeated measures ANOVA. For the ALDQ, differences between new and existing hearing aid users were examined using Mann-Whitney U tests. Additionally, whether ALDQ responses were associated with seven-week follow-up outcomes (GHAB/DiffP, HHIE, VFS-AHL, DOSO, BKB-SIN, hearing aid datalogging) was investigated using Spearman’s rank (non-parametric) correlation coefficient.

#### Non-validated self-report measures

For the CCQ and Hearing Tasks Diary, differences between new and existing hearing aid users were examined using Mann-Whitney U tests. For the smartphone app feedback survey, common themes were generated by AH from participants’ responses to open-ended questions using an established thematic analysis procedure, which comprises specific analytical phases: data familiarization, generating initial codes, searching for themes, reviewing themes, and defining and naming themes (Braun & Clarke, 2006). The analysis was inductive and themes were defined as something important about the data that represented repeated patterns of response or meaning that were prevalent (i.e. reported by several participants) across the entire data set. To ensure that the interpretation of the data was not limited to the perspective of one individual, a second author (RG) independently coded responses and formulated potential themes. Any discrepancies were discussed amongst AH and RG, and an agreement was made regarding which themes should be applied.

#### Behavioural measures

For the BKB SIN test, both performance and perceived listening effort data were skewed. As a result, differences between test conditions at the one- and seven-week follow-up study sessions were compared using separate Mann-Whitney U tests. Differences between sessions for each test condition were examined using Wilcoxon signed-rank tests. For hearing aid datalogging, the difference between the one- and seven-week follow-up sessions for new and existing hearing aid users was assessed using separate paired samples t-tests.

Bonferroni correction for multiple comparisons was applied for each outcome measure. Effect sizes, small (≤ .2), medium (≤ .5) and large (≤ .8) (Cohen, 1988), and 95% confidence intervals (CIs) for group differences are reported. Statistical significance was set to *p*< .05.

## RESULTS

### Study Population

Forty-four eligible participants consented to take part in the study between September 27^th^ 2018 and January 11^th^ 2019. Demographic information across the entire sample, as well as for new and existing hearing aid users, is provided in Supplemental Materials. Overall, participants were representative of a typical UK clinical population, presenting with mild to moderate hearing loss (*mean pure-tone threshold in the better ear across 0.25-4 kHz*= 38.5 dB HL; *SD*= 8.66), a mean age of 68.8 years (*SD*= 9.52), a mean MoCA score of 26.3 (*SD*= 1.95), and most (75%) self-reported as ‘*competent*’ users of digital technologies. Existing hearing aid users had owned conventional hearing aids for an average of 8.8 years (*SD*= 6.23). Figure 1 provides a flow diagram showing progression of participants through each phase of the study. Prior to the baseline visit, one new hearing aid user withdrew, citing that they no longer had time to continue participation in the study due to family commitments. After the baseline session, two existing hearing aid users declined to participate further; one was dissatisfied with the smartphone-connected hearing aids and the other was unhappy about returning the devices on completion of the study.

### Outcome Measures

#### Validated self-report measures

Mean subscale scores for each outcome measure are provided separately for new and existing hearing aid users in Supplemental Materials.

##### Glasgow Hearing Aid Benefit Profile (GHABP)

On average, new hearing aid users reported that they experienced *moderate* hearing disability (52.6%) and handicap (54.7%) at baseline (i.e., unaided). In comparison, following seven-weeks of using smartphone-connected hearing aids, self-reported disability significantly decreased (i.e. improved) to 11.6%, with a large effect size, *t*(11)= 7.67, *p*< .001, *d*= 2.95 (Figure 2A). In addition, on average new users reported that they used the smartphone-connected hearing aid *all of the time* (use, 100%), the *hearing aid was a great help* or *hearing is perfect with the aid* (benefit, 80.7%), and were *very satisfied* or *delighted with the aid* (satisfaction, 84.4%).

**Figure 2.**
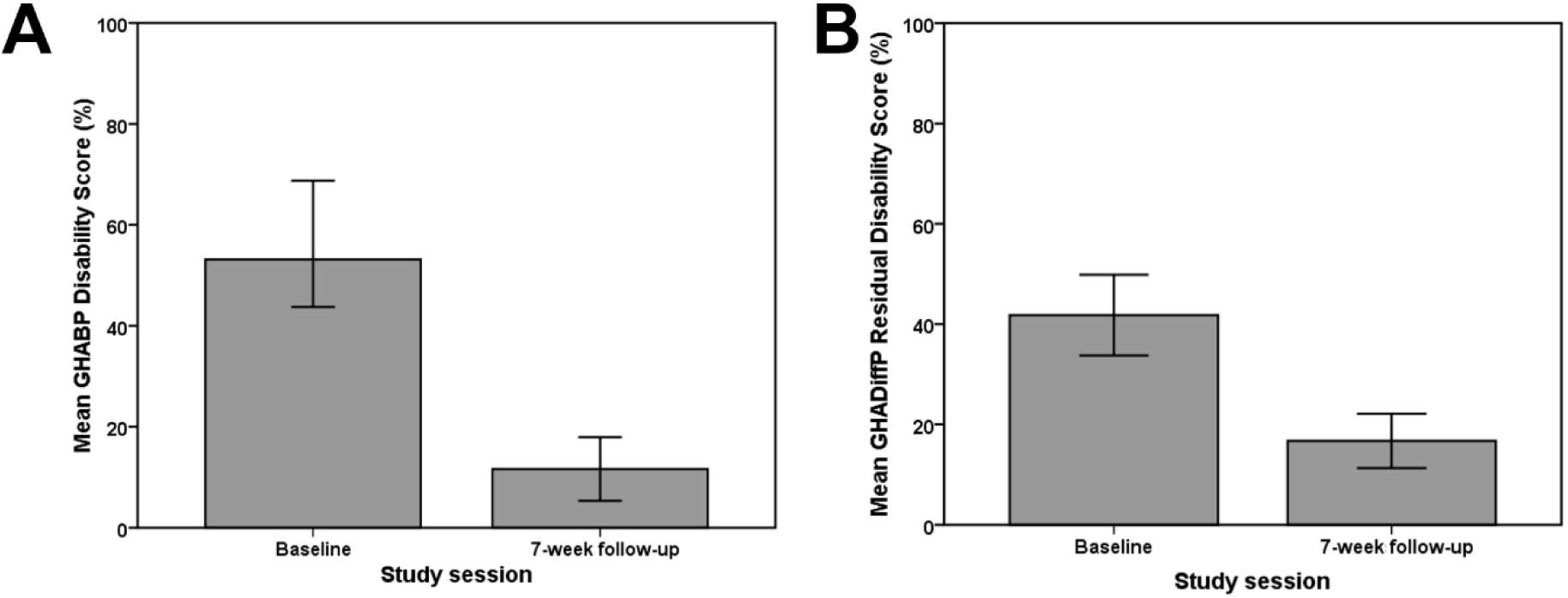
Mean disability subscale scores for (A) new hearing aid users completing the Glasgow Hearing Aid Benefit Profile (GHABP), and (B) existing hearing aid uses completing the Glasgow Hearing Aid Difference Profile (GHADiffP), at baseline and following seven-weeks of using the smartphone-connected hearing aids. Error bars=95% CI.

##### Glasgow Hearing Aid Difference Profile (GHADiffP)

At baseline, existing hearing aid users reported, on average, that they used their conventional hearing aids *all the time* (use, 100%) and experienced *only slight/moderate difficultly* (residual disability, 41.8%%). Following seven-weeks of using the smartphone-connected hearing aids, self-reported use did not differ statistically from that reported for conventional hearing aids at baseline (100%, *p*= .168), likely due to ceiling effects. However, self-reported residual disability significantly reduced (i.e. improved) to 16.7% for the smartphone-connected compared to the conventional hearing aids, *t*(28)= 6.83, *p*< .001, *d*= 1.39 (Figure 2B). Additionally, compared to the conventional hearing aids, the smartphone-connected hearing aids were reported as *much better* (benefit, 100%), and users were *much more satisfied* (satisfaction, 100%) with them.

##### Hearing Handicap Inventory for the Elderly (HHIE)

For both new and existing hearing aid users, overall scores significantly reduced (i.e., improved) from baseline (new: *M*= 44.67, *SD*= 18.42; existing: *M*= 36.41, *SD*= 25.06) to the seven-week follow-up (*M*= 7.17, *SD*= 6.35; *M*= 19.52, *SD*= 15.86), *t*(11)= 6.13, *p*< .001, *d*= 1.77, and *t*(28)= 3.31, *p*= .003, *d*= .62, respectively. A similar pattern of results was also found for emotional and social/situational subscales (Figure 3A).

**Figure 3.**
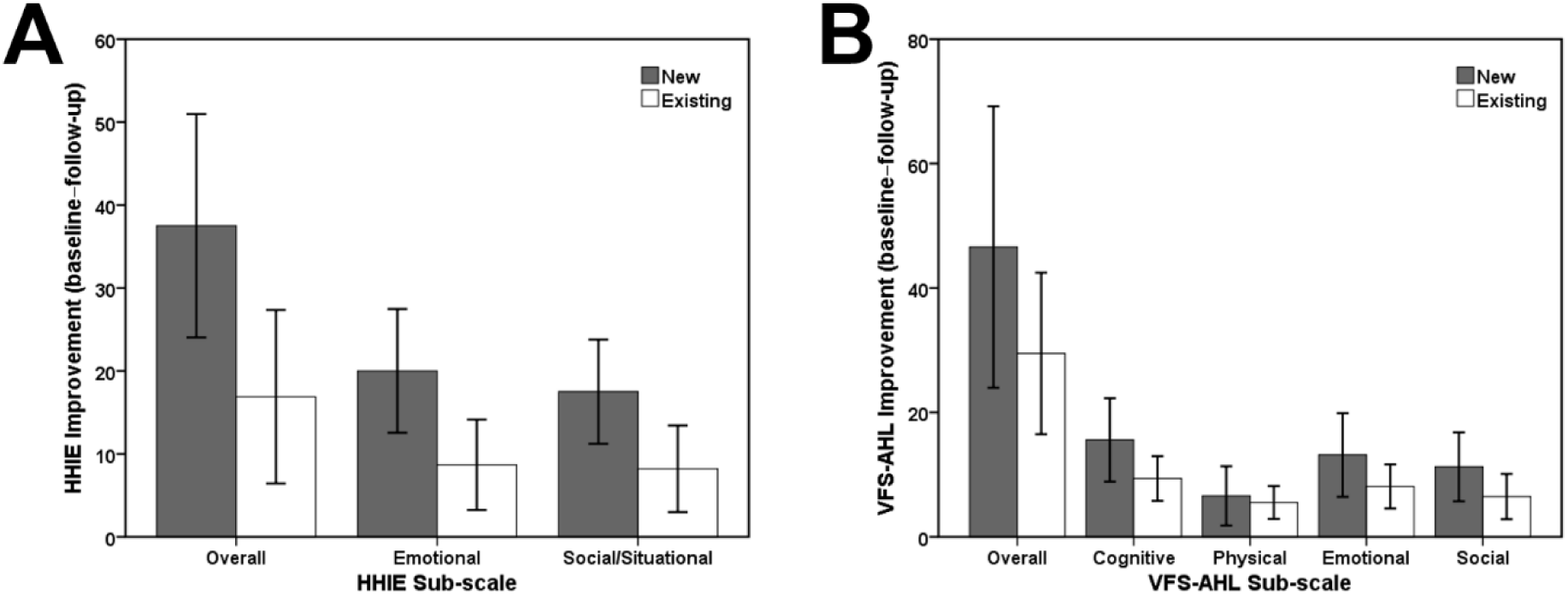
Mean improvement (i.e., difference between baseline and seven-week follow-up) for (A) Hearing-Handicap Inventory for the Elderly (HHIE), and (B) Vanderbilt Fatigue Scale for Adults with Hearing Loss (VFS-AHL) reported by new and existing hearing aid users. Error bars=95% CI.

##### Vanderbilt Fatigue Scale for Adults with Hearing Loss (VFS-AHL)

For both new and existing hearing aid users, overall listening-related fatigue scores significantly reduced (i.e., improved) from baseline (*M*= 60.17, *SD*= 31.85; *M*= 59.17, *SD*= 37.47) to the seven-week follow-up (*M*= 13.58, *SD*= 11.67; *M*= 29.69, *SD*= 28.21), *t*(11)= 4.53, *p*< .001, *d*= 1.31, and *t*(28)= 4.65, *p*< .001, *d*= .86, respectively. A similar pattern of results was also found for cognitive, physical, emotional, and social subscales (Figure 3B).

##### Device Orientated Subjective Outcome Scale (DOSO)

For existing users, scores for the speech cues (baseline: *M*= 3.66, *SD*= .95; one-week: *M*= 5.45, *SD*= .71; seven-week: *M* =5.56, , *SD*= 1.02), listening effort (*M*= 4.52, *SD*= 1.00; *M*= 6.10, *SD*= .48; *M*= 6.17, *SD*= .83), pleasantness (*M*= 4.58, *SD*= 1.03; *M*= 5.23, *SD*= .93; *M*= 5.61, *SD*= 1.25), quietness (*M*= 3.11, *SD*= 1.16; *M*= 5.58, *SD*= .85; *M*= 5.71, *SD*= 1.14), and convenience (*M*= 5.02, *SD*= .88; *M*= 5.94, *SD*= .80; *M*= 6.06, *SD*= .98) subscales were all significantly better for smartphone-connected hearing aids compared to conventional hearing aids (i.e. baseline) when assessed at the one- (*p*≤ .001 , *d*≥ .93) and seven-week follow-up (*p*≤ .005, *d*≥ .87) study sessions (Figure 4). In comparison, there was no differences between self-reported use for the conventional hearing aids assessed at baseline (*M*= 4.53, *SD*= .80) and the smartphone-connected hearing aids assessed at the one- (*M*= 4.77, *SD*= .51) and seven-week (*M*= 4.68, *SD*= .62) follow-up study sessions (*p*≥ .480).

**Figure 4.**
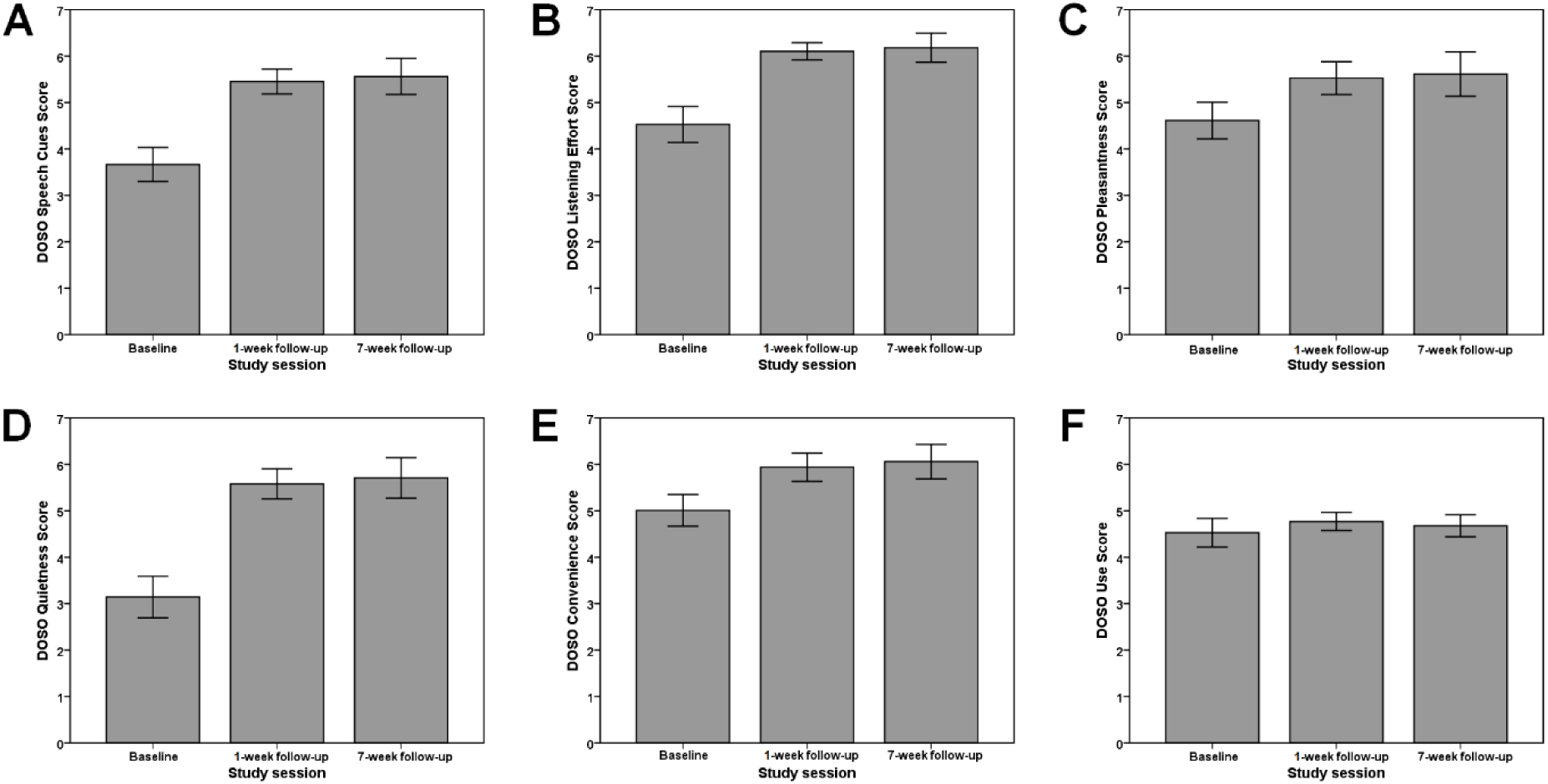
Mean Device Orientated Subjective Outcome Scale (DOSO) subscale scores for (A) speech cues, (B) listening effort, (C) pleasantness, (D) quietness, (E) convenience, and (F) use, reported by existing hearing aid uses for conventional hearing aids (baseline) and smartphone-connected hearing aids (one- and seven-week follow-up). Error bars=95% CI.

##### Auditory Lifestyle and Demands Questionnaire (ALDQ)

The median responses for each question at the one-week follow-up session are shown in Supplemental Materials. Responses did not differ statistically between new and existing hearing aid users (*p*≥ .944). Both user groups reported that, on average, they *Sometimes* (*Med*= 2, *IQR*= 0) find themselves in difficult listening situations and that this was of *Some Importance* (*Med*= 2, *IQR*= 1) in their everyday lives. Furthermore, ALDQ scores were not significantly associated with any outcome measure scores assessed at the seven-week follow-up (*p*≥ .546).

#### Non-validated self-report measures

##### Initial Hearing Aid Preferences Questionnaire (IHAPQ)

Responses did not differ statistically between new and existing hearing aid users (*p*≥ .530). Therefore, the median responses obtained for each question at baseline in rank order across both user groups are shown in Supplemental Materials. The highest-ranking preference was that participants wanted hearing aids to be customised to meet their needs by a hearing healthcare professional (*Med*= 10, *IQR*= 2). This was closely followed by preferences that participants were able to optimise their hearings themselves using smartphone technologies (*Med*= 9, *IQR*= 2), and should be allowed to program and customise the hearing aids themselves (*Med*= 9, *IQR*= 3). The least important preferences (although still with a relatively high median responses) were for hearing aids to work completely automatically ‘out-of-the-box’ (*Med*= 8, *IQR*= 4), be small and hardly visible (*Med*= 8, *IQR*= 4), and help to manage tinnitus (*Med*= 8, *IQR*= 5).

##### Clarity and Comfort Questionnaire (CCQ)

As there were no statistically significant differences between individual preferences for new and existing hearing aid users (*p*≥ .343), the results are reported collectively for both groups (Supplemental Materials). When wearing hearing aids, most participants reported that they prefer sounds to be *clear and sharp* (70.7%) compared to *comfortable and soft* (29.3%). In addition, when listening at a lower volume, participants preferred *listening at that volume* if they can still hear well (73.2%) rather than at a *slightly higher volume* (26.8%) and are *only sometimes* easily disturbed by unpleasantly loud sounds (90.2%). Responses to whether participants *find sudden sounds in the listening environment unpleasantly loud* were approximately equal (Yes= 56.1%; No= 43.9%).

##### Hearing Tasks Diary

As shown in Supplemental Materials, for new hearing aid users, gain setting preferences varied considerably depending on the pre-set and listening situation. By comparison, existing users consistently reported that the gain setting *‘on’ was slightly better* for every pre-set and listening situation specified, except for the *more sound* gain setting used with the *TV* pre-set when *watching a film*. Nevertheless, preference ratings only differed statistically between new and existing hearing aid user groups for the *clarity* gain setting used with the *restaurant* pre-set when listening in a *restaurant when talking to one or several other people* (*U*= 43.5, *p*= .016). In this situation, the median preference for new users was that *clarity ‘off’ was much better* (*Med*= 2, *IQR*= 2), whereas existing users expressed that *clarity ‘on’ was slightly better* (*Med*= -1, *IQR*= 1.5).

##### Feedback survey

As responses did not differ statistically between new and existing hearing aid users (*p*≥ .132), results are reported collectively in Supplemental Materials. For the closed-ended questions, most participants reported that the app met their needs either *very well* (36.8%) or *extremely well* (31.6%). The app was rated highly (*M*= 3.92 out of 5 stars, *SD*= .82), and the situation that the app was deemed most useful was having a conversation in a noisy environment (50%), followed by watching the TV (31.6%). In addition, the hearing aid setting (or modifier) that participants liked most was volume (31.6%) or noise reduction (31.6%), whereas the least liked setting was soft/loud (5.3%) or mid gain (2.6%). For the open-ended questions, it was consistently stated that a key benefit of the app was the ability to make personalised adjustments to the hearing aid settings (40.5%), as well as being able to use the app in any listening situation (32.4%). When asked, *What changes would this app have to make you give it a higher rating?*, 27.6% of participants responded *nothing*.

However, a high proportion of participants (34.5%) would have liked the app to allow even greater adjustment and personalisation, as well as improved reliability of the Bluetooth connection between the app and hearing aids (18.7%). In terms of their general experiences with the hearing aids, over half (57.1%) of participants reported that the sound-quality provided by the smartphone-connected hearing aids was superior compared to their previous, conventional hearing aids. Only 4.8% commented that the smartphone-connected hearing aids were no different to their existing devices. In addition, 16.7% of participants stated that the accompanying smartphone app was beneficial, particularly in noisy listening situations, compared to 7.1% who stated that it was not. A small number (9.5%) of participants stated that the hearing aids were uncomfortable to wear due to the cShell couplings.

#### Behavioural measures

##### BKB Speech-In-Noise Test

There were no significant differences between new and existing hearing aid users for performance (*p*≥ .148) or effort ratings (*p*≥ .274) across all factory pre-set each test conditions (*restaurant/neutral, restaurant/clarity, custom*). Therefore, results are reported collectively for both hearing aid user groups in Supplemental Materials. The proportion of correct keywords identified was high across test conditions at both one- (restaurant/neutral: *Med*= 96, *IQR*= 5; restaurant/clarity: *Med*= 96, *IQR*= 6.75; custom: *Med*= 94.5, *IQR*= 4.75) and seven-week follow-up sessions (*Med*= 96, *IQR*= 4; *Med*= 97, *IQR*= 4; *Med*= 95, *IQR*= 4.75), approaching ceiling. Although performance was slightly lower for the custom hearing aid pre-set test condition relative to both restaurant/neutral and restaurant/clarity, these differences were not statistically significant at either the one- (*p*≥ .573) or seven-week follow-up (*p*≥ .195) study sessions. In addition, for each test condition, performance did not statistically differ between the one- and seven-week follow-up sessions (*p*≥ .285). A similar pattern of results was also found for perceived listening effort ratings, whereby test conditions were consistently rated as ‘*little effort*’ at the one- (*Med*= 4, *IQR*= 3; *Med*= 4, *IQR*= 4; *Med*= 5, *IQR*= 4) and seven-week follow-up sessions (*Med*= 4, *IQR*= 4; *Med*= 4, *IQR*= 4; *Med*= 5, *IQR*= 3). As such, there were no significant differences between test conditions at the one- (*p*≥ .387) and seven-week follow-up sessions (*p*≥ .129), or between the one- and seven-week follow-up sessions (*p*≥ .405) for each test condition.

##### Hearing aid use (datalogging)

For both new and existing hearing aid users, average hours of daily use for smartphone-connected hearing aids recorded at the one-week follow-up session were higher (new: *M*= 12.28 hours, *SD*= 3.5; existing: *M*= 12.10, *SD*= 3.44) compared to that at the seven-week follow-up session (*M*= 11.38, *SD*= 3.40; *M*= 11.73, *SD*= 3.58). While this difference was not statistically significant for new users (*p*=.242), it was for existing users, *t*(26)= 3.73, *p*< .001, *d*= .72 (Figure 5).

**Figure 5.**
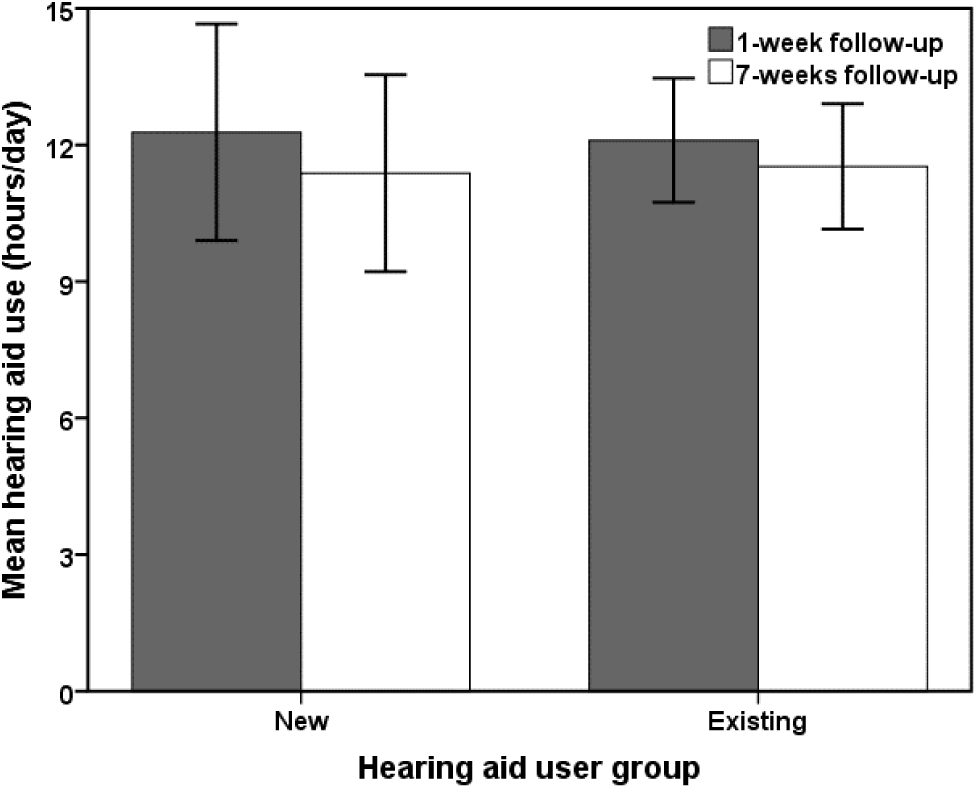
Mean use (hours per day) for the smartphone-connected hearing aids recorded using datalogging at the one- and seven-week follow-up sessions for new and existing hearing aid users. Error bars= 95% CI.

## DISCUSSION

Increasing rates of smartphone ownership in older adults suggest that there is a growing demand for the delivery of health services via mobile technologies, termed mHealth (Cucciniello et al., 2021). In the context of hearing healthcare, mHealth is expanding rapidly as an accessible and affordable method of delivering patient-centred care to facilitate hearing loss management (Ferguson, 2019; Ferguson, Maidment, Henshaw, & Heffernan, 2019). Arguably, one of the most prominent and increasingly available mHealth interventions in adult aural rehabilitation is smartphone-connected hearing aids. However, evidence assessing these devices has been limited (Maidment et al., 2018). To address this need, the current study quantitatively evaluated the benefits, or otherwise, of using smartphone-connected hearing aids and an accompanying app in both new and existing hearing aid users.

Overall, we found that smartphone-connected hearing aids improved self-reported outcomes irrespective of prior hearing aid experience, all with moderate-to-large clinical effect sizes. In terms of hearing-related difficulties, activity limitations (disability) and participation restrictions (handicap) significantly improved following seven-weeks of using the smartphone-connected hearing aids. These findings support those found in previous qualitative studies assessing the usability of smartphone-connected hearing aids in existing users only (Gomez et al., 2022; Maidment et al., 2019; Ng et al., 2017). Specifically, previous research has shown that smartphone-connected hearing aids can increase social participation, a finding that has been attributed to the user’s ability to make fine-tune adjustments to their hearing aid settings via an accompanying app.

The current study also found that hearing loss-related fatigue across multiple dimensions (cognitive, physical, emotional, social) improved in both user groups. Previous research has shown that adults living with hearing loss often report high levels of hearing loss-related fatigue in their everyday lives, which has been attributed to increased listening effort and concentration (Alhanbali et al., 2017; Holman et al., 2019; Hornsby et al., 2016). While amplification might reduce effortful listening in adults with hearing loss, there is limited evidence evaluating the effects of hearing aid fitting on fatigue (Holman et al., 2021). This study may help to address this evidence gap, suggesting that smartphone-connected hearing aids can reduce hearing loss-related fatigue in both new and existing users. Even so, further research is warranted to investigate the impact of smartphone connectivity on fatigue. This is pertinent given the findings of our companion qualitative study (Gomez et al., 2022), which demonstrated that there may be an additional cognitive burden associated with making adjustments via an app whilst simultaneously trying to communicate in difficult and rapidly changing listening situations.

With regards to hearing aid outcomes, self-reported use, benefit, and satisfaction for smartphone-connected hearing aids were all high in both new and existing hearing aid users. For existing users, quality of listening across a range of domains (i.e., speech cues, listening effort, pleasantness, quietness, convenience) were all superior for smartphone-connected devices relative to their previous conventional hearing aids. This latter finding could be attributable to the additional functionalities provided by the accompanying smartphone app, including greater opportunities to conveniently adjust and customise the hearing aid programmes and algorithms. However, as this was an observational study, additional research, such as a randomised controlled trial (RCT), is needed to definitively test this.

Despite this, the results of our purpose-designed feedback questionnaire found that the app met users’ listening and communication needs *extremely* or *very well* in 68% of cases, and increased opportunities for them to tailor their hearing aid settings in any listening situation. Moreover, participants consistently reported that user-centred adjustment was one of the most beneficial aspects of the app, especially in difficult listening situations, such as when having a conversation in the presence of background noise. These quantitative findings are further supported by our qualitative study (Gomez et al., 2022), where it was found that smartphone-connected hearing aids led to feelings of empowerment, whereby self-tuning via the app facilitated problem solving, coping mechanisms, and use of hearing aids. A similar pattern of results was also found by Maidment et al. (2019), whereby adjusting hearing aids via an app provided users with a greater sense of autonomy and control, empowering them to successfully manage their hearing loss in everyday life. More recently, the concept of empowerment across the aural rehabilitation journey has been explored, with five key dimensions identified – knowledge, participation, strategies and skills, self-efficacy, and control (Gotowiec et al., 2022). Critically, all dimensions are represented in the outcomes from the current study and in our companion qualitative paper, confirming the concept of empowerment as a key benefit of smartphone-connected hearing aids. Therefore, we recommend that empowerment should be assessed across all five dimensions using appropriate outcome measures (e.g., Bennett et al., in review; Gotowiec et al., in review) to illuminate how smartphone-connected hearing aids, as well as other technologies, can empower adults to manage their hearing loss.

In addition to self-tuning via the app, it should be acknowledged that improved hearing aid outcomes may be attributable to the featural differences between the smartphone-connected hearing aids assessed, which may have been more technologically advanced compared to participants’ previous conventional NHS hearing aids. Namely, in this study participants trialled Phonak Audeo B90-Direct smartphone compatible hearing aids, whereas most existing users had previously been using Phonak Nathos S+ Micro devices. The Phonak Audeo B90-Direct has 20 fine-tuning compression channels compared to 16 for the Phonak Nathos S+ Micro. Furthermore, while both hearing aids have an adaptive multichannel directional microphone, the Phonak Audeo B90-Direct includes additional automatic and manual programs, such as wind and comfort in echo, which are not available for the Phonak Nathos S+ Micro. Differences in available features between devices may subsequently explain why existing users’ subjective ratings of the smartphone-connected hearing aids were higher compared to their conventional hearing aids. Featural difference may also explain why a high proportion of participants commented via the feedback survey that the sound-quality provided by the smartphone-connected hearing aids was superior. Therefore, an RCT comparing the same smartphone-connected hearing aids with and without the app would be necessary to determine the extent to which self-adjustment via this method leads to further, incremental benefits.

Taken together, the current findings, as well as those reported elsewhere (Gomez et al., 2022; Maidment et al., 2019; Maidment & Ferguson, 2018; Ng et al., 2017), add to a growing body of evidence suggesting that smartphone-connected hearing aids benefit adults living with hearing loss. When coupled with research showing that audiologists are generally supportive of incorporating smartphone-connected hearing aids into their aural rehabilitation practices (Kimball et al., 2018; Ng et al., 2017; Olson et al., 2022), it is becoming increasingly apparent that smartphones have the potential to transform hearing healthcare delivery. Thus, based on the available research evidence to-date, we argue that audiologists should consider smartphone-connected hearing aids for all those who have access to smartphone, and ensure that the functionality of the accompanying smartphone app is (i) enabled, and (ii) explained to the new user. Given our research, and that of others, to not do so is a disservice to hearing aid users, particularly older adults. We consider that, by using suitable behaviour change techniques, audiologists can support their patients to use these new technologies (Gomez et al., 2022). Furthermore, by reframing the smartphone as a tool to remain connected with family, friends, and the wider world, we can move away from a practitioner-centred (or ‘paternalistic’) mode of hearing healthcare, to one where individuals are empowered to be in control of their hearing and hearing aids.

## AKNOWLEDGMENTS

This research was funded co-funded by the NIHR Nottingham Biomedical Research Centre and Sonova AG and carried out at/supported by the NIHR Nottingham Clinical Research Facilities. The views expressed are those of the authors and not necessarily those of the NHS, the NIHR or the Department of Health and Social Care. We would like to thank Paige Church for assisting with participant recruitment and data collection.

## Data Availability

All data produced in the present study are available upon reasonable request to the authors.

## Supplemental Materials

**Supplemental Materials 1.** Median response for new and existing hearing aid users for each of the 24 listening situations assessed in the Auditory Lifestyle and Demands Questionnaire (ALDQ). For each situation, participants are asked how often (i.e., frequency) they find themselves in that situation (1 = *very rarely*, 2 = *sometimes*, or 3 = *often*), as well as how important it is for them to be able to hear in that situation in their everyday life (1 = *very little*, 2 = *some importance*, or 3 = *very important*).

| Question | New |  | Existing |  |
| --- | --- | --- | --- | --- |
|  | *Frequency* | *Importance* | *Frequency* | *Importance* |
| Listening in a background of noise. | 2.5 (1) | 2.5 (1) | 2 (1) | 3 (1) |
| Listening to sounds that are quiet and difficult to hear. | 2 (1) | 2 (1) | 2 (1) | 3 (1) |
| Listening to sounds that are consistently loud. | 1.5 (1.75) | 1.5 (1) | 2 (1) | 2 (1) |
| Listening to things that are close by you. | 3 (.75) | 3 (0) | 3 (1) | 3 (1) |
| Listening to things that are far away. | 2 (1) | 2 (1.75) | 2 (1) | 2 (1) |
| Listening when there are lots of echoes. | 1 (1.75) | 1 (1) | 1 (1) | 1 (1.25) |
| Listening when two or more people are talking at once. | 3 (0) | 3 (1) | 3 (1) | 3 (1) |
| Having to listen to sounds that vary a lot in loudness. | 2 (1.75) | 2 (1) | 2 (0) | 2 (.25) |
| Talking on the telephone. | 3 (.75) | 3 (0) | 3 (1) | 3 (0) |
| Listening to music at home. | 2.5 (1) | 2.5 (1) | 2 (1) | 2 (1) |
| Listening to music at a concert. | 2 (1) | 2 (2) | 2 (1) | 2 (1) |
| Listening to the radio. | 3 (0) | 3 (1) | 2.5 (1) | 2 (1) |
| Listening to the television. | 3 (0) | 3 (0) | 3 (0) | 3 (1) |
| Listening to sounds or voices that are moving around. | 2 (.75) | 2 (.75) | 2 (1) | 2 (1) |
| Listening when you have no control or influence over the speaker. | 2 (.75) | 2 (1.75) | 2 (.25) | 2 (1) |
| Listening when not being able to understand could be embarrassing. | 2 (1) | 3 (0) | 2 (1) | 3 (1) |
| Listening when misunderstanding could cause an accident. | 1 (0) | 2.5 (2) | 1 (1) | 3 (1.25) |
| Listening when misunderstanding could cause you to lose money. | 1 (0) | 3 (2) | 1 (0) | 2 (2) |
| Listening to a speaker who is in another room. | 1 (1) | 2 (1) | 2 (2) | 2 (2) |
| Listening to a speaker whose voice is not familiar to you. | 2 (0) | 2 (0) | 2 (1) | 2 (0) |
| Listening to the sounds of nature. | 2 (1.5) | 2 (1.75) | 3 (1) | 2.5 (1) |
| Having to understand what is happening around you. | 3 (.75) | 3 (0) | 2.5 (1) | 3 (1) |
| Listening to people with unfamiliar accents or dialects. | 2 (0) | 2.5 (1) | 2 (0) | 2 (1) |
| Being in a situation where you do not want to hear what is happening. | 2 (1) | 2.5 (1) | 2 (1) | 2 (1) |

*Brackets indicate IQR.*

**Supplemental Materials 2.** Median responses for each question on the Initial Hearing Aid Preferences Questionnaire (IHAPQ) completed at baseline. Questions are provided in rank order (highest to lowest preference).

| **Question** | **Response** |
| --- | --- |
| Hearing aids should be customised to my needs by a HHCP. | 10 (2) |
| I should be able to optimise the hearing aid settings myself using my smartphone. | 9 (2) |
| I should be able to use and wear hearing aids all day regardless of what I am doing. | 9 (2) |
| Hearing aids should be robust, resist moisture, sweat, dust, rain and are waterproof. | 9 (3) |
| Hearing aids should allow me to listen to sounds directly from electronic devices. | 9 (3) |
| I should be allowed to program and customise the hearing aids myself. | 9 (3) |
| Hearing aids should work completely automatically, once programmed by a HHCP. | 9 (3) |
| Hearing aids should work completely automatically 'out of the box'. | 8 (4) |
| Hearing aids should be very small and hardly visible when worn. | 8 (4) |
| Hearing aids should help me manage tinnitus. | 8 (5) |

*Brackets indicate IQR.*
*HHCP, hearing healthcare professional.*

**Supplemental Materials 3.** Number and percentage of responses for each question on the Clarity and Comfort Questionnaire (CCQ) assessing individual preferences regarding sound quality from hearing aids.

| **When I wear my hearing aids:** | **N (%)** |
| --- | --- |
| I prefer sound to be... |  |
| ...*clear and sharp* | 29 (70.7) |
| ...*comfortable and soft* | 12 (29.3) |
| If I can hear well at a lower volume, I prefer... |  |
| ...*that volume* | 30 (73.2) |
| ...*volume slightly higher* | 11 (26.8) |
| I am easily disturbed by sounds around me. |  |
| ...*Only sometimes* | 37 (90.2) |
| ...*Yes* | 4 (9.8) |
| I find that sudden sounds in the listening environment are unpleasantly loud. |  |
| ...*Yes* | 23 (56.1) |
| ...*No* | 18 (43.9) |

**Supplemental Materials 4.** Median gain settings preferred for new and existing hearing aid users for each factory pre-set within the app in specified listening situations, as measured by the Hearing Tasks Diary assessing different. Preferences rated on a five-point scale (−2= *gain setting ‘on’ sounds much better*; -1=, *‘on’ sounds slightly better*; 0=, *‘on’ sounds the same as ‘off’*; 1=, *‘off’ sounds slightly better*; 2= *‘off’ sounds much better*).

| **Pre-set** | **Gain setting** | **Listening situation** | **New** | **Existing** |
| --- | --- | --- | --- | --- |
| TV | More sound | Watching a film | -0.5 (2) | 0 (0.5) |
| TV | Clarity | Watching the news | 0 (1.5) | -1 (2) |
| Restaurant | Clarity | At a Restaurant talking to one or several other people | 2 (2) | -1 (1.5) |
| Restaurant | Comfort | In a noisy social gathering | -2 (1) | -1 (2.5) |
| Music | More base | Listening to music | -0.5 (2) | -1 (2) |
| Music | More treble | Listening to music | 0.5 (1) | -1 (1.5) |
| Car | Clarity | Passenger in a car talking to driver | 0 (3) | -1 (0.8) |
| Car | Comfort | Passenger in a car listening to traffic noise | -1 (2) | -1 (2) |

*Brackets indicate IQR.*

**Supplemental Materials 5.** Participant responses to the purpose-designed feedback survey.

*Brackets indicate IQR.* *Supplemental Materials 5. Participant responses to the purpose-designed feedback survey.*
| **Question** | **Response options** | **(%)** |
| --- | --- | --- |
| How has your experience been with the hearing aids, in general?* | Improved sound quality compared to previous hearing aids | 57.1 |
|  | No different to previous hearing aids | 4.8 |
|  | Accompanying app beneficial | 16.7 |
|  | Accompanying app not beneficial | 7.1 |
|  | Improved hearing noticed by others | 4.8 |
|  | Uncomfortable to wear | 9.5 |
| What does the app do well?* | Self-adjustment/personalisation | 40.5 |
|  | Use in different listening situations | 32.4 |
|  | Empowerment | 18.9 |
|  | Automatic changes | 5.5 |
|  | Easy to use | 2.7 |
| What changes would this app have to make you give it a higher rating?* | More personalisation | 34.5 |
|  | Nothing | 27.6 |
|  | Improved Bluetooth connectivity | 13.8 |
|  | More noticeable change to sound settings | 10.3 |
|  | Simplify controls within the app | 6.9 |
|  | Would prefer to make changes using a different device (e.g., remote control, button on hearing aid) | 6.9 |
| Overall, how well does the app meet your needs? | Extremely well | 31.6 |
|  | Very well | 36.8 |
|  | Somewhat well | 26.3 |
|  | Not so well | 2.6 |
|  | Not at all well | 2.6 |
| How would you rate the app? | Five stars | 26.3 |
|  | Four stars | 42.1 |
|  | Three stars | 28.9 |
|  | Two stars | 2.6 |
|  | One star | 0 |
| Which setting (or modifier) did you like most? | Volume | 31.6 |
|  | Noise reduction | 31.6 |
|  | Base | 10.5 |
|  | Treble | 10.5 |
|  | Sound focus | 7.9 |
|  | Soft/loud | 5.3 |
|  | Mid | 2.6 |
| In which situation is the app most useful? | Conversation in a noisy environment | 50 |
|  | Watching TV | 31.6 |
|  | Conversation with 1-2 people | 5.3 |
|  | In the office/during work | 7.9 |
|  | In the car | 2.6 |
|  | Listening to music | 0 |

\* Open-ended question

**Supplemental Materials 6.** Demographic information of consenting participants.

| Demographic variable | Hearing aid user group |  |  |
| --- | --- | --- | --- |
|  | New | Existing | Overall |
| **Gender (N)** |  |  |  |
| Female | 4 (28.6%) | 9 (30%) | 13 (29.5%) |
| Male | 10 (71.4%) | 21 (70%) | 31 (70.5%) |
| **Age (years)** |  |  |  |
| Mean | 66.71 | 69.8 | 68.82 |
| SD | 11.21 | 8.66 | 9.52 |
| **Five frequency<sub>0.25-4kHz</sub> better ear PTA (dB HL)** |  |  |  |
| Mean | 35.5 | 39.83 | 38.45 |
| SD | 9.05 | 8.36 | 8.66 |
| **Montreal Cognitive Assessment (MoCA)** |  |  |  |
| Mean | 26.5 | 26.17 | 26.27 |
| SD | 1.78 | 2.04 | 1.95 |
| **Digital technology competency (N)** |  |  |  |
| Beginner | 1 (7.2%) | 5 (16.7%) | 6 (13.6%) |
| Competent | 10 (71.4%) | 23 (76.7%) | 33 (75.0%) |
| Did not specify | 3 (21.4%) | 2 (6.6%) | 5 (11.4%) |

**Supplemental Materials 7.** Mean outcome measures for new hearing aid users (n=14) at baseline, one-week and/or seven-weeks follow-up study sessions.

| Outcome measure | Baseline |  | 1-week |  | 7-week |  |
| --- | --- | --- | --- | --- | --- | --- |
| GHABP (*new only*) |  |  |  |  |  |  |
| Disability | 52.6 | (17.16) | - | - | - | - |
| Handicap | 54.69 | (21.67) | - | - | - | - |
| Use* | - | - | - | - | 100 | (15.15) |
| Residual disability | - | - | - | - | 11.63 | (9.91) |
| Benefit | - | - | - | - | 80.73 | (14.95) |
| Satisfaction* | - | - | - | - | 84.38 | (25.00) |
| HHIE |  |  |  |  |  |  |
| Overall | 44.67 | (18.42) | - | - | 7.17 | (6.35) |
| Emotional | 23.33 | (9.88) | - | - | 3.33 | (3.94) |
| Social/situational | 21.33 | (8.79) | - | - | 3.83 | (3.46) |
| DOSO |  |  |  |  |  |  |
| Speech cues | - | - | 5.6 | (0.78) | 5.76 | (0.69) |
| Listening effort | - | - | 5.99 | (0.79) | 6.25 | (0.60) |
| Pleasantness | - | - | 4.83 | (1.49) | 5.5 | (1.06) |
| Quietness | - | - | 4.83 | (1.42) | 5.45 | (1.07) |
| Convenience | - | - | 5.67 | (1.06) | 5.83 | (0.76) |
| Use | - | - | 4.58 | (0.61) | 4.56 | (0.52) |
| VFS-AHL |  |  |  |  |  |  |
| Overall | 60.17 | (31.85) | - | - | 13.58 | (11.67) |
| Cognitive | 21.58 | (8.63) | - | - | 6 | (4.65) |
| Physical | 8.33 | (7.99) | - | - | 1.75 | (1.71) |
| Emotional | 15.92 | (9.42) | - | - | 2.75 | (3.36) |
| Social | 14.33 | (7.82) | - | - | 3.08 | (3.90) |
| Hearing aid datalogging | - | - | 12.28 | (3.75) | 11.38 | (3.40) |

\* Median
Brackets indicate SD or IQR
GHABP, Glasgow Hearing Aid Benefit Profile; GHADiffP, Glasgow Hearing Aid Difference Profile; HHIE, Hearing Handicap Inventory for the Elderly; VFS-AHL, Vanderbilt Fatigue Scale for Adults with Hearing Loss; DOSO, Device Orientated Subjective Outcome Scale

**Supplemental Materials 8.** Mean outcome measures for existing hearing aid users (n=30) at baseline, one-week and/or seven-week follow-up study sessions.

| Outcome measure | Baseline |  | 1-week |  | 7-week |  |
| --- | --- | --- | --- | --- | --- | --- |
| GHADiffP |  |  |  |  |  |  |
| Use ('old' aids)* | 100 | (13.54) | - | - | - | - |
| Residual disability ('old' aids) | 41.81 | (21.15) | - | - | - | - |
| Use ('new' aids)* | - | - | - | - | 100 | (6.25) |
| Residual disability ('new' aids) | - | - | - | - | 16.74 | (14.22) |
| Benefit ('old' vs. 'new' aids)* | - | - | - | - | 100 | (12.50) |
| Satisfaction ('old' vs. 'new' aids)* | - | - | - | - | 100 | (15.63) |
| HHIE |  |  |  |  |  |  |
| Overall | 36.41 | (25.06) | - | - | 19.52 | (15.86) |
| Emotional | 18 | (14.65) | - | - | 9.31 | (8.80) |
| Social/situational | 18.41 | (10.97) | - | - | 10.21 | (8.56) |
| VFS-AHL |  |  |  |  |  |  |
| Overall | 59.17 | (37.47) | - | - | 29.69 | (28.21) |
| Cognitive | 19.56 | (9.78) | - | - | 10.28 | (8.69) |
| Physical | 10.1 | (8.08) | - | - | 4.57 | (5.75) |
| Emotional | 15.14 | (10.16) | - | - | 7.03 | (6.87) |
| Social | 14.28 | (10.53) | - | - | 7.79 | (7.95) |
| DOSO |  |  |  |  |  |  |
| Speech cues | 3.66 | (0.95) | 5.45 | (0.71) | 5.56 | (1.02) |
| Listening effort | 4.52 | (1.00) | 6.10 | (0.48) | 6.17 | (0.83) |
| Pleasantness | 4.58 | (1.03) | 5.23 | (0.93) | 5.61 | (1.25) |
| Quietness | 3.11 | (1.16) | 5.58 | (0.85) | 5.71 | (1.14) |
| Convenience | 5.02 | (0.88) | 5.94 | (0.8) | 6.06 | (0.98) |
| Use | 4.53 | (0.80) | 4.77 | (0.51) | 4.68 | (0.62) |
| Hearing aid datalogging (hours/day) | - | - | 12.1 | (3.44) | 11.53 | (3.48) |

\* Median
Brackets indicate SD or IQR
GHABP, Glasgow Hearing Aid Benefit Profile; GHADiffP, Glasgow Hearing Aid Difference Profile; HHIE, Hearing Handicap Inventory for the Elderly; VFS-AHL, Vanderbilt Fatigue Scale for Adults with Hearing Loss; DOSO, Device Orientated Subjective Outcome Scale

**Supplemental Materials 9.** Median percent correct keywords identified during the BKB-SIN test for restaurant/neutral, restaurant/clarity, and custom hearing aid factory pre-set test conditions. Median perceived listening effort, rated on a visual analogue scale ranging from one (‘*No Effort*’) to 13 (‘*Extreme Effort*’), were also provided by participants on completion of each test condition.

| **Test condition** | **1-week** |  | **7-week** |  |
| --- | --- | --- | --- | --- |
|  | % Correct | Effort | % Correct | Effort |
| Restaurant/neutral | 96 (5) | 4 (3) | 96 (4) | 4 (4) |
| Restaurant/clarity | 96 (6.75) | 4 (4) | 97 (4) | 4 (4) |
| Custom | 94.5 (4.75) | 5 (4) | 95 (4.75) | 5 (3) |

*Brackets indicate IQR*

